# Machine learning-based personalized composite score dissects risk and protective factors for cognitive and motor function in elderly

**DOI:** 10.1101/2022.11.18.22282498

**Authors:** Ann-Kathrin Schalkamp, Stefanie Lerche, Isabel Wurster, Benjamin Roeben, Milan Zimmermann, Franca Fries, Anna-Katharina von Thaler, Gerhard Eschweiler, Thomas Gasser, Walter Maetzler, Daniela Berg, Kathrin Brockmann, Fabian H. Sinz

**Author notes:** Corresponding author: Fabian Sinz PhD & Kathrin Brockmann MD, Institut für Informatik, University of Göttingen, Goldschmidtstraße 1, 37077 Göttingen, Germany, Department of Neurodegeneration, Hertie Institute for Clinical Brain Research, University of Tuebingen, Hoppe Seyler-Strasse 3, 72076 Tuebingen, Germany. These Authors contributed equally (Ann-Kathrin Schalkamp, Stefanie Lerche). These Authors contributed equally and share senior authorship (Kathrin Brockmann, Fabian H. Sinz).

## Abstract

Most nations worldwide have aging populations. With age, sensory, cognitive and motor abilities decline and the risk for neurodegenerative disorders increases. These multiple impairments influence the quality of life and increase the need for care, thus putting a high burden on society, the economy, and the healthcare system. Therefore, it is important to identify factors that influence healthy aging, in particular ones that are potentially modifiable by each subject through choice of lifestyle. However, large-scale studies that investigate the influence of multiple multi-modal factors on a global description of healthy aging measured by multiple clinical assessments are sparse. Here, we propose a Machine Learning (ML) model that simultaneously predicts multiple cognitive and motor outcome measurements on a personalized level recorded from one learned composite score. This personalized composite score is derived by the model from a large set of multi-modal components from the TREND cohort including genetic, biofluid, clinical, demographic and lifestyle factors. We found that a model based on a single composite score was able to predict cognitive and motor abilities almost as well as a flexible regression model specifically trained for each single clinical score. In contrast to the flexible regression model, our composite score-based model is able to identify factors that globally influence cognitive and motoric abilities as measured by multiple clinical scores. We used the model to identify several risk and protective factors for healthy aging and recover physical exercise as a major, modifiable, protective factor.

## Introduction

Neuropsychiatric diseases are currently the leading cause of disability and dependency worldwide. Among them, the neurodegenerative diseases Parkinson’s disease (PD) and Alzheimer’s dementia (AD) are rising fastest ^1^. Aging represents one of the strongest risk factors for both diseases and predictions indicate that the number of cases will double worldwide in the next 20 years as the proportion of elderly steadily increases ^2^. Thus, the identification and effect size of risk and protective factors that indicate the dynamic processes from aging to accelerated aging and potentially to neurodegeneration with motor and cognitive dysfunction are of high interest for aging societies. While some factors such as sex and genetic status are immutable, a large proportion can be influenced by lifestyle. These factors include arterial hypertension, adiposity, diabetes mellitus, smoking, physical activity, muscle mass, and education. This offers the opportunity to focus on these factors for preventive health care strategies. However, many of these factors are interdependent. Moreover, the heterogeneity of human subjects and the intervals at which genetic, biochemical, clinical and demographic markers are collected in longitudinal studies make it difficult to extract suitable data for robust statistical predictions. Thus, conventional statistical analyses with pair-wise, step-wise or even multivariate analyses might not adequately reflect effect sizes of each single factor and their combined effects. To overcome these limitations, we used an unbiased machine learning (ML) approach on our large longitudinal TREND cohort (Tübingen Risk Evaluation for Neurodegenerative Diseases) comprising 1200 elderly participants aged between 50-80 and followed-up for up to 10 years. We developed a Bayesian model to simultaneously predict aging-related key functions, namely motor and cognitive performance, on an individual level from a single composite score that reflects a large set of multi-modal factors including genetic, biofluid, clinical, demographic and lifestyle factors.

Similar models have been used in high-dimensional medical settings using imaging or genetic data ^34^ and also to investigate dietary patterns by fat types ^4^. However, such methods have not been used in multi-modal settings assessing aging- and neurodegeneration-related profiles. Specifically, we employed a Reduced Rank Regression (RRR) approach (see Figure 1) in order to identify whether one composite score can predict various age-related abilities on an individual level. The higher the composite score, the worse the performance across age-related key functions. By determining whether a factor has a positive or negative impact on the composite score, we were able to classify it as a risk-increasing or protective factor. Furthermore, we used a monotonic transformation to model ordinal predictors which allowed us to gain knowledge about the relationship between the categories of an ordinal variable, such as identifying recessive or dominant effects of genetic markers. Importantly, we primarily focused on factors that were already identified in epidemiological and genetic studies by standard statistical approaches in order to facilitate a proof-of-concept for the Bayesian model and compared the predictive performance of the Bayesian model to a standard statistical approach with multiple regression modeling for each outcome measures. We found that – our single composite score can predict multiple age-related domains of motor and cognitive function. The main drivers of our composite score were hours of exercise and muscle mass to be among the most protective mutable factors while higher body mass index was identified as a risk factor. Furthermore, our model indicates that hours of exercise has a proportional effect instead of a binary effect in the sense that more exercise also has a more protective effect.

**Figure 1:**
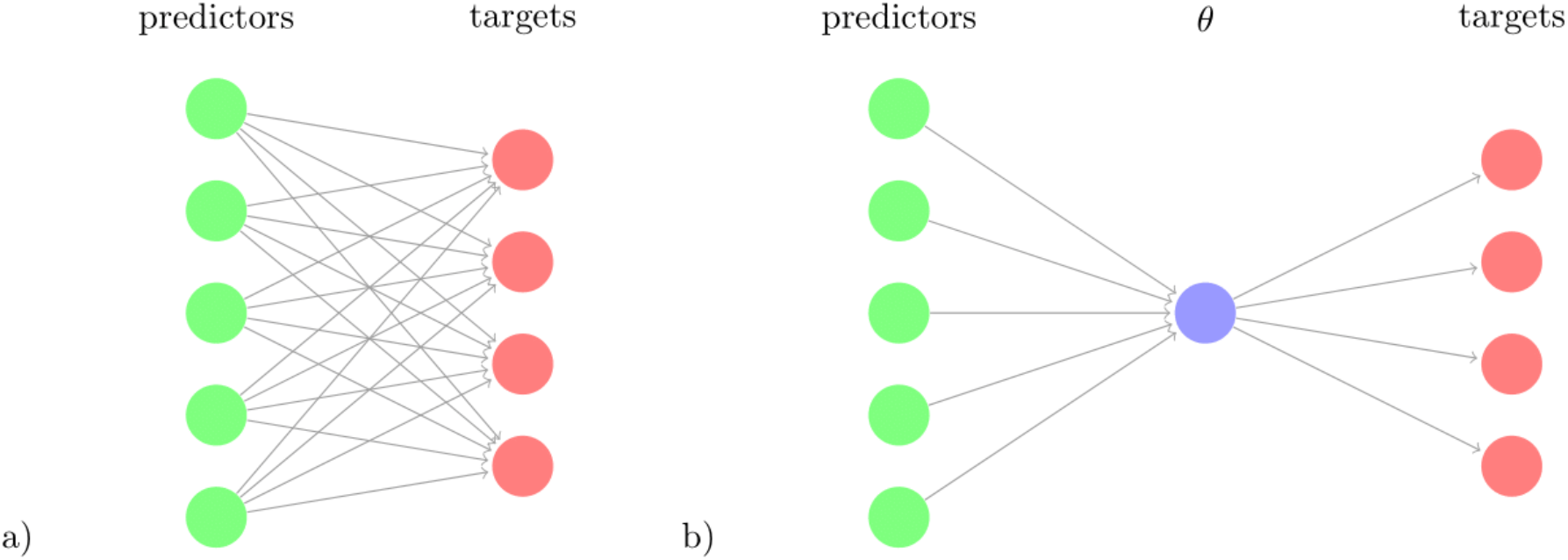
Schematic comparison of multivariate regression and reduced rank regression. The amount of coefficients to learn (number of arrows) is illustrated for a) multivariate regression and b) reduced rank regression. The latent variable θ, the composite risk, is a linear combination of the predictors and is projected via linear multiplications to the targets.

## Results

### Model Performance

The Bayesian RRR composite score model achieved a comparable performance to classical linear regressions (OLS) per clinical outcome measure (Figure 2). It showed similar levels of explained variance for all cognitive outcome measures (Supplementary Table 3). Separate linear regressions significantly outperformed the composite score model in only four outcome measures (Walk while Cross Dual – Single, Subtract while Walk Dual – Single, Walk while Subtract Dual + Single, Walk while Cross Dual + Single). All of these are gait-related outcome measures. Importantly, our composite score model performed on par with linear regressions in predicting the gait-related outcome measure “Subtract while Walk Dual + Single’’ which is the gait measure that has the highest cognitive load as it measures the speed of mathematical calculations while walking. Overall, the composite score model performed well on cognitive outcome measures, on par with the OLS models and worse on gait-related measures. This indicates that a composite score RRR model with a single explaining factor performs almost equally well as several individual OLS models.

**Figure 2:**
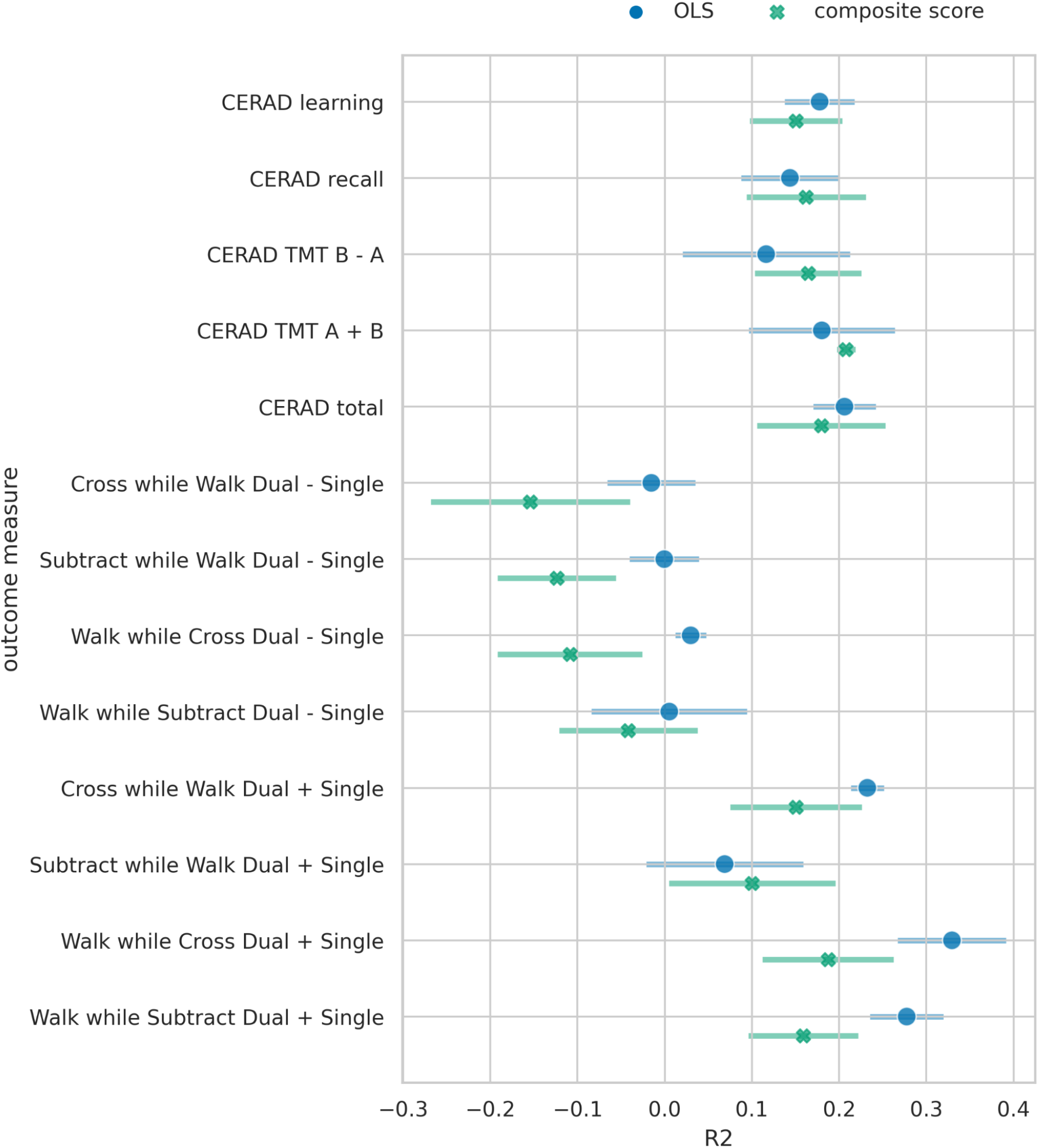
Performance comparison of Bayesian RRR and multiple OLS. The mean 5-fold CV R2 on the test sets is shown for each of the outcome measures. Error bars denote the 95% Confidence Interval across five folds. For the Bayesian RRR we show the mean across 5 folds of the mean R2 over 1000 samples and the corresponding 95% Confidence Interval across 5 folds.

To assess whether the worse performance in the four gait-related measures is due to the reduced complexity of our model or due to other model specifications, we trained a separate Bayesian RRR for each outcome measure. These performed as well as the OLS models on all tasks (see A6).

### All models recover known protective and risk factors

#### Multiple linear regressions (OLS)

The multiple linear regressions showed an overall agreement for the effect direction, i.e. whether a factor is a risk or a protective factor (Figure 3). Factors identified as protective are female sex, longer time in education, and higher level of physical fitness (hours of exercise per week, higher skeleton muscle mass). Only for some movement speed-related outcome measures (Walk while Subtract/Cross Dual - Single) female sex negatively impacted the outcome measure (i.e. decreased the performance). Risk factors that were in agreement between the majority of the OLS models are older age, higher number of cigarette pack-years, and a higher BMI.

**Figure 3:**
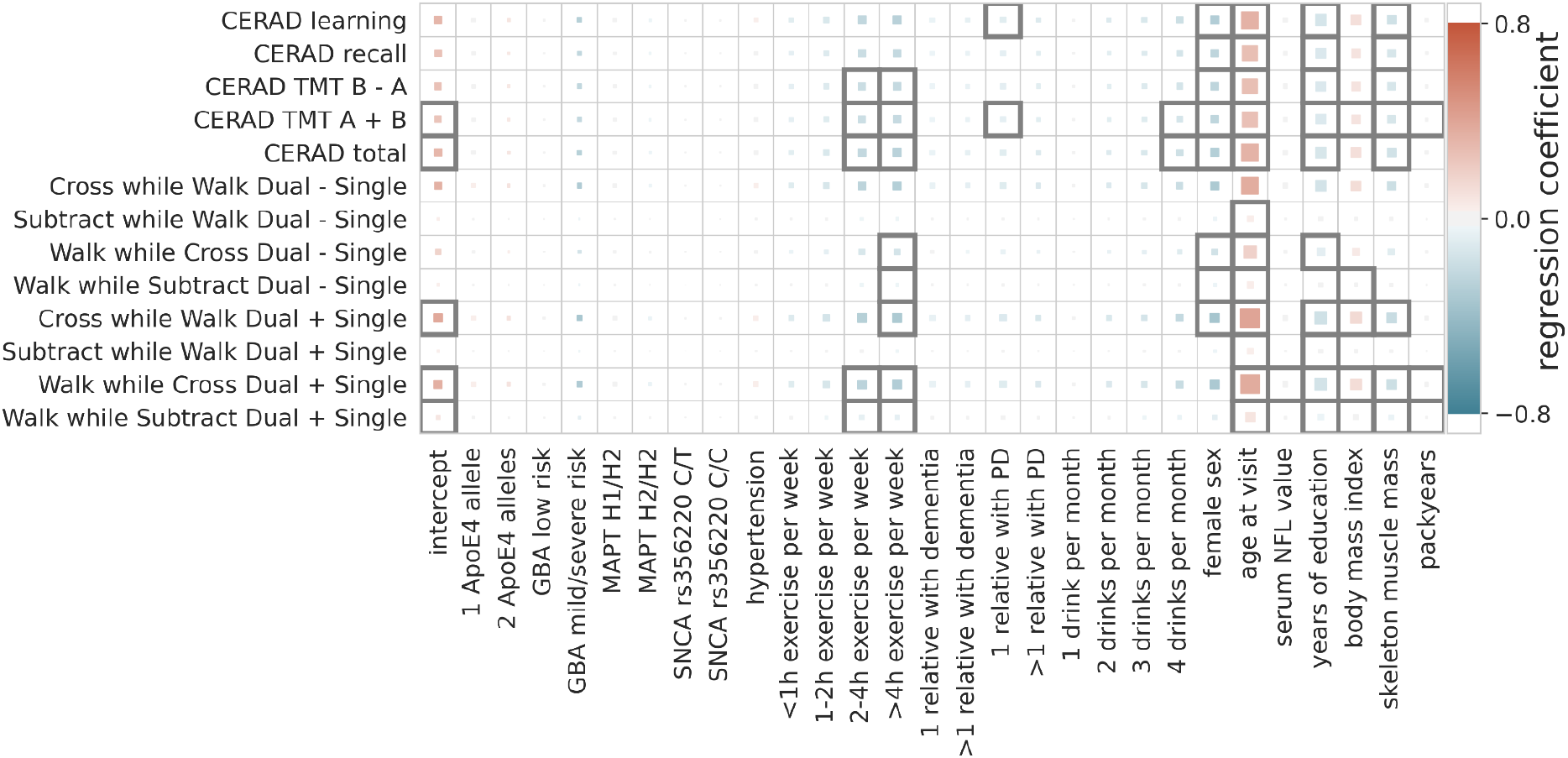
*Regression coefficients of multiple linear models. The influence of the predictive factors (x-axis) onto the outcome measures (y-axis) is shown. The color indicates the size and direction of the effect (protective = blue, risk = red) with the size showing the importance (abs(coefficient)/standard error) and a black outline indicating significance (Bonferroni-corrected p-threshold 0*.*05)*.

#### Bayesian Reduced Rank Regression composite score model

The composite score Bayesian reduced rank regression model merged this overall agreement of the OLS models into one composite score (Figure 4 and Figure 5). In addition to the identified factors from the OLS models the Bayesian model identified hypertension, *ApoE4* genotype, and higher NFL values as significant risk factors. The number of relatives with PD or dementia were not significant in any OLS model but were identified as significant protective factors in our composite risk model. The Bayesian RRR further identified genetic variants in *GBA* and *MAPT (H2 haplotype)* as protective factors.

**Figure 4:**
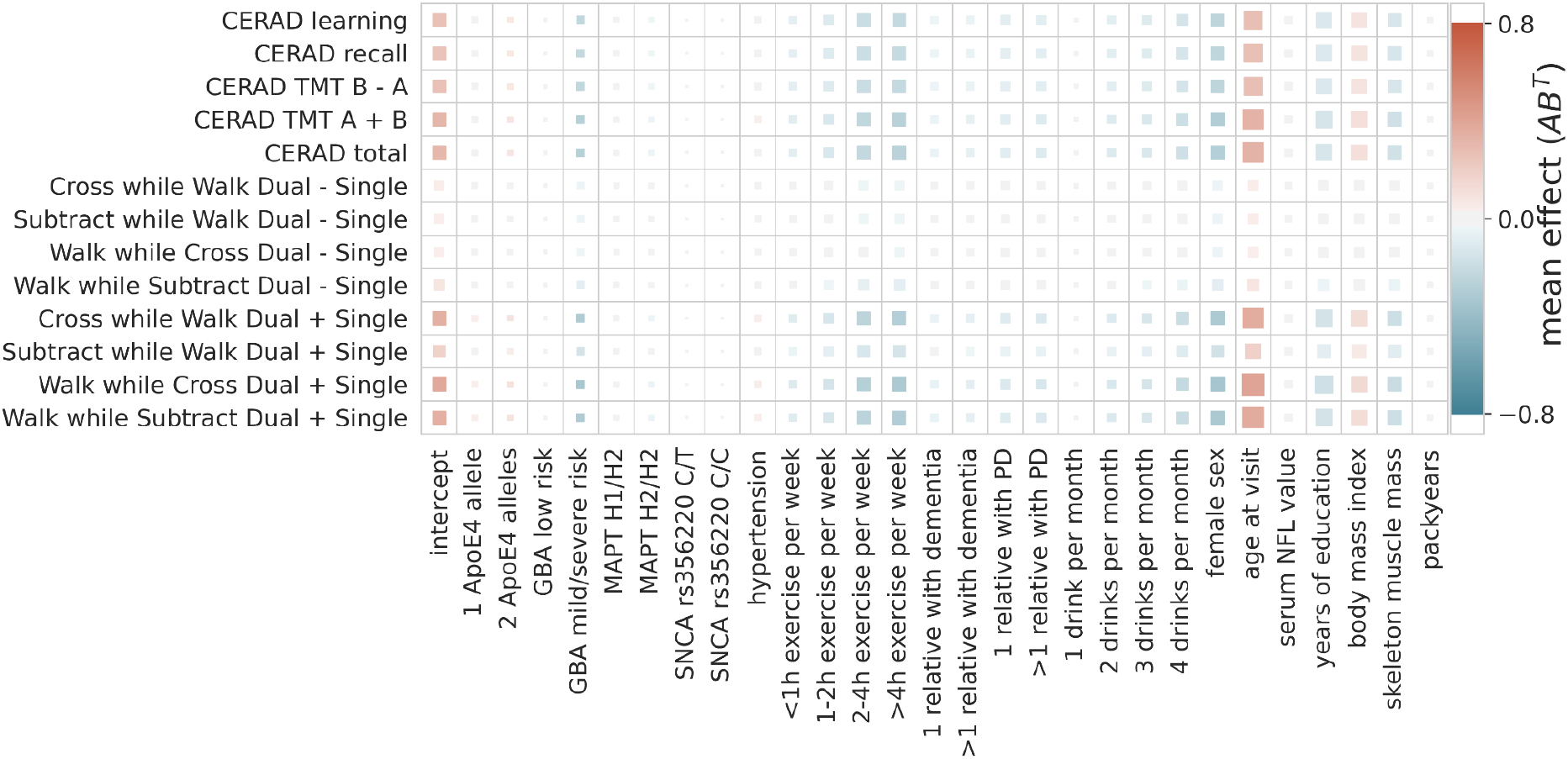
The composite score model recovers the overall agreement of the coefficients across the multiple OLS models. The color indicates the direction and size of the effect of a predictor (x-axis) onto a target (y-axis). The size of the square indicates its importance as the absolute ratio of mean and standard deviation (the larger the further away from 0).

**Figure 5:**
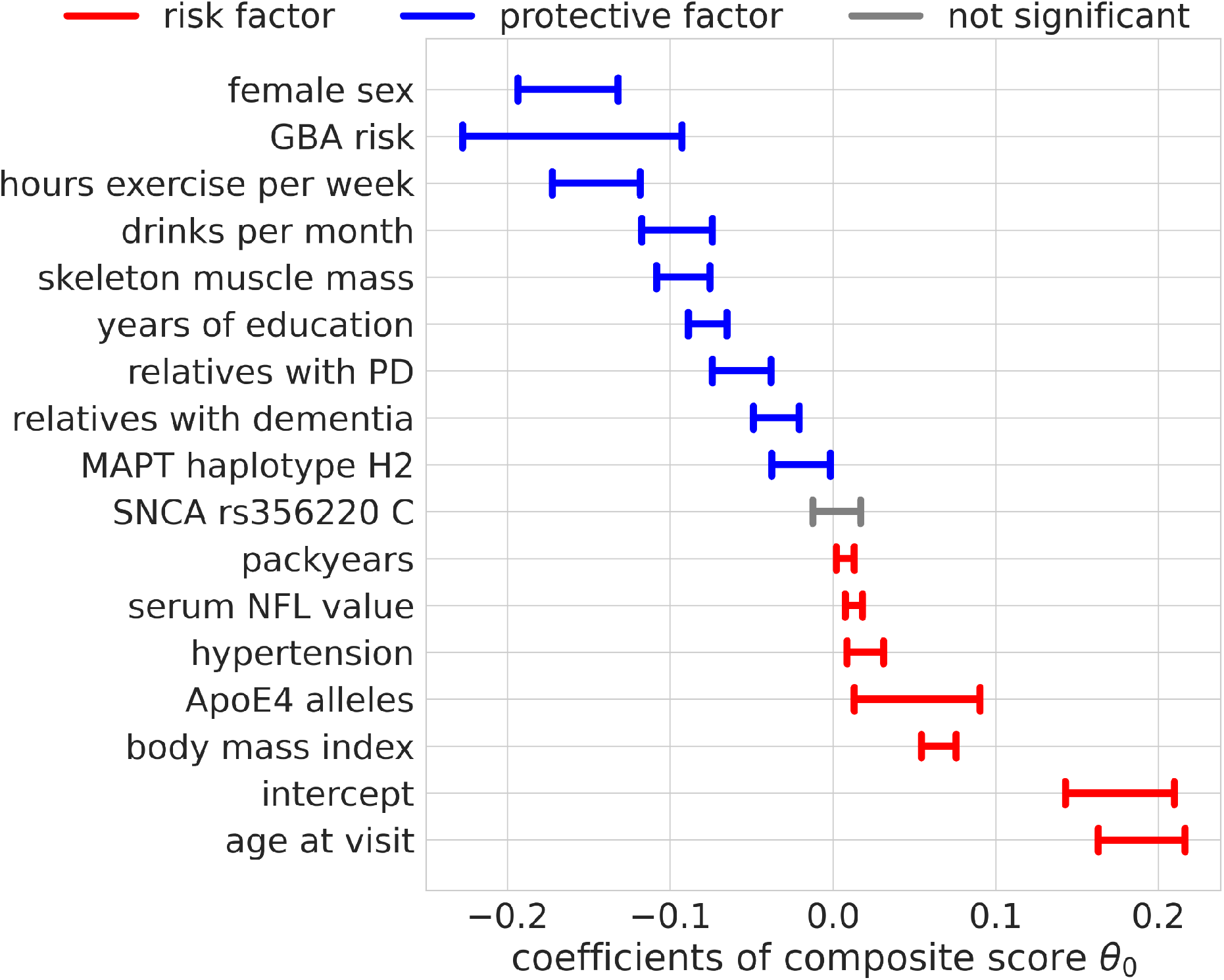
Regression coefficients for the composite score model. The estimated effect sizes of the predictors on the composite score are displayed. The highest posterior density is plotted. The coloring indicates significance (95% highest posterior density contains 0, not significant = gray) and direction of the effect (blue = protective, red = risk).

#### Monotonic transformation reveals a proportional effect for exercise

The encoding of ordinal factors in our model allowed for fine grained information on their effect onto the composite score not addressed by a nominal encoding as used in the OLS models. By learning the distance between the categories of ordinal predictors through monotonic transformation we obtained flexible spacing of the different levels with additional meaning (Figure 6). We saw a steep reduction of risk for people who drink at least 2 drinks per month but increasing the number of drinks did not further reduce the risk substantially. In contrast, for physical exercise we observed no such saturation and can conclude that more exercise is more protective. We also note a steep increase in risk for carriers of 2 *ApoE4* alleles compared to carriers of one allele. Contradictorily, heterozygous carriers of mild and severe *GBA* variants seem to be more protected than those with *GBA* wildtype.

**Figure 6:**
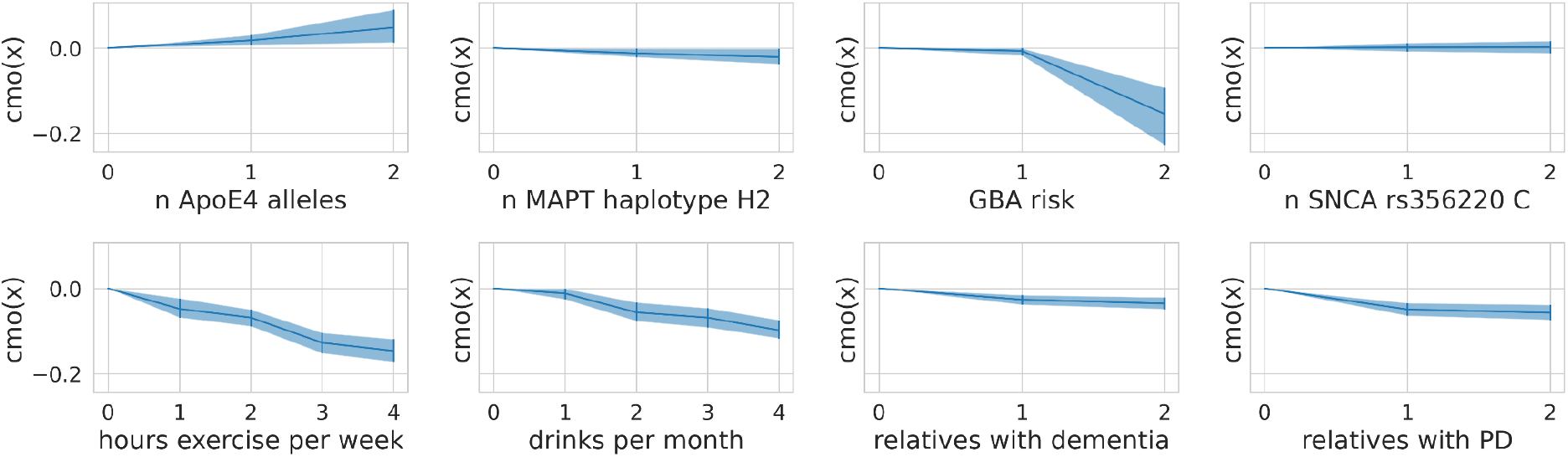
Flexible spacing of ordinal predictors. For each ordinal predictor we show the distance between the categories modeled through monotonic transformation in the composite score model. We plot the learned distances multiplied by the predictors effect size (A). The mean of the samples alongside the highest posterior density interval (95%) is shown.

## Discussion

We analyzed the combined influence of a large set of multi-modal factors, including environmental, lifestyle, biofluid and genetic data, on aging-related key functions, cognitive and gait performance, measured by multiple clinical tests. To this end, we compared two approaches: independent prediction of each outcome measure with a linear regression model (OLS) and joint prediction of all outcome measures from one composite score learned by a Bayesian reduced rank regression model. We could show that the predictive performance of the Bayesian RRR model with one single composite score was comparable to classical multiple OLS models. The most relevant factors that showed a protective effect on complex gait and cognitive abilities in elderly were female sex as well as a higher degree of physical activity, more skeleton muscle mass, and more years of education. Higher age, higher body mass index and more smoking pack-years, the presence of hypertension, having two ApoE4 alleles, and higher serum levels of NFL were predictors for impaired gait and reduced cognitive performance. However, note that our model does not test for causality but merely identifies associations between the included factors and outcome measures. Statements about whether increasing physical activity will improve cognitive and motor functioning cannot be made without further causal assumptions on the involved factors.

We primarily included well known risk and protective factors in order to check the validity of the composite modeling. However, some factors showed an influence in an unexpected direction. For example, the number of relatives with dementia or PD should serve as a proxy of the genetic risk and thus be a risk factor. Our model as well as the OLS models instead revealed a protective effect. This could be due to a high motivation of individuals with a family member affected by a neurodegenerative disease as they have an increased personal interest in performing well and taking care of one’s health. Such motivational influences were not measured directly within the TREND study but literature supports this assumption^5^. Carriers of mild and severe variants in the *GBA* gene had a reduced risk according to our model which could be a reflection of these carriers being significantly younger than the other groups (mild and severe *GBA* variant carriers (N = 30) vs wildtype (N = 4294): t-statistic = - 3.12, p-value = 1.8e-3; mild and severe *GBA* variant carriers (N = 30) vs low risk *GBA* variant carriers (N = 166): t-statistic = -2.54, p-value = 1.2e-2).

Our modeling of the ordinal predictors allowed for interpretations of the effect sizes of each category. For example, *ApoE4* is a well-known genetic risk factor for cognitive decline with carriers of one allele having an odds ratio of about 3 for developing AD and carriers of two alleles having an odds ratio of about 15^6^. This steep increase in risk for carriers of two alleles was replicated in our model despite the small sample size (11 persons with two ApoE4 alleles and 193 with one *ApoE4* allele). In general, the sample size of carriers of specific genetic variants (*GBA, SNCA, MAPT*) probably did not suffice to recover their effect on age-related functions.

We investigated how much complexity is needed to achieve similar performance to the OLS models in all outcome measures by training a model with two composite scores and by training separate Bayesian models for each outcome measure. Increasing the latent space and allowing for two composite scores slightly improved the performance for a subset of the clinical tests, albeit not significantly. This mainly affected gait outcome measures, and revealed distinct effects of factors for different clinical tests (Supplementary Table 3). For example, female sex was identified as a risk factor for walking speed in general but a protective factor across all cognitive tests (Figure 3). This effect was found in the OLS models and the two composite scores model (Supplemental Figure 3 and 4) alike and could reflect the height difference and thus step size differences between males and females. This was not measured within the TREND study and can thus not be corrected for. The single composite score model thus prioritized the cognitive measures over the gait-related measures leading to a composite score that performs well on cognitive measures and worse than the OLS models on the gait measures. The good performance of the single composite score model to predict the gait measure “Subtract while Walk Dual + Single” might be explained through the high cognitive load of this task. It might thus be better represented by a cognitive composite score.

Our approach of a joint prediction of all outcome measures from one composite score learned by a Bayesian RRR model performed comparably well to the more classical flexible individually fitted regression models. This suggests that already one composite score can capture a substantial part of the complex effects on cognitive and motor function in an aging cohort. Our model unbiasedly identified known risk and protective factors of aging.

The fact that our less flexible model performed comparably well to individual OLS models indicates that the explored factors either share similar mechanistic pathways and/or are inter-related to each other. This further highlights a global underlying risk for aging processes where motor and cognitive abilities are affected alike.

Our Bayesian RRR has several strengths compared to traditional approaches such as its handling of missing data, its reduced complexity and thus its interpretability, and its handling of ordinal predictors. In Bayesian models, missing values in the outcome measures can be imputed through the model’s parameter estimates. As we can include incomplete data into the model, we increased the total amount of data the model uses but did not alter the data distribution artificially by learning from imputed outcome measures (covariates were imputed). Through our assumption of a composite risk, we decreased the complexity of the model and thus made it more scalable and better suited for medical data, which are scarce and high-dimensional. This assumption of a low rank further increased the model’s interpretability as the single composite score can be interpreted as an estimate of the true underlying risk. The modeling of the ordinal predictors better captured the true scale level of the data indicating that physical exercise has an additive effect.

Our model currently assumes a linear relation between the predictors and outcomes although this is not necessarily true. For example, it would be reasonable to assume that drinking a small amount of alcohol could have a protective effect but excessive drinking could be a risk factor for cognitive performance. Such reverse effects cannot be captured through our linear models. Using a quadratic link function could possibly better account for such scenarios, other non-linearities could be further explored through neural networks. Drinking two drinks might be a confounding fitness factor, as many seniors avoid drinking if they are multimorbid or take multiple medications.

Another limitation of our current model is the handling of longitudinal data, where we treat visits from the same individual as independent. This disregards the correlation within a subject. A potential improvement of our model could be the adaptation of a mixed model where visits are grouped by individuals and identified through a subject specific identifier, i.e. random effects. This would further allow us to make statements about a subject’s temporal slope. A different approach to model the longitudinal data would be a stacked model where a linear mixed model first learns the trajectory over time for each subject and this estimated change over time is then used as the outcome measure in our Bayesian RRR.

Currently, our model achieved similar performance to multiple OLS models, however, several adaptations could be explored to improve our model’s performance. As our model requires less parameters, we could increase the amount of predictors and targets without the need to increase the sample size. Especially exploring the effect of various aging-related genetic markers and their interaction could be a promising future project. A similar model has been used before for modeling genotype-phenotype associations ^3^ however, they did not use monotonic transformations but instead assumed additive effects for the SNPs.

We conclude that our low parametric modeling approach successfully recovered known risk and protective factors of healthy aging on a personalized level while providing an interpretable composite score. An extension of this model using more predictors and clinical tests could further identify unknown factors and distinct aging-related processes. To this end, more sensitive tests are needed that better capture the variation with a healthy cohort. Digital sensors such as wrist-worn acceleration devices could provide such sensitive data. The modeling approach is generalizable and could also be applied to other cohorts to investigate the complex interplay of risk and protective factors along with effect sizes from different dimensions such as lifestyle, medical, genetic and biochemical data.

## Methods

### Study Population

We used the data from the TREND study (Tuebingen Risk Evaluation for Neurodegenerative Diseases) which is a prospective longitudinal study initiated in 2009 with biennial assessments of elderly participants aged between 50 and 80 years without neurodegenerative diseases at study recruitment. Newspaper announcements and public events were used to recruit participants from Tübingen and the surrounding area. Between 2009 and 2012, 1201 participants underwent baseline assessments. For study inclusion, participants had to be free of a diagnosis of a neurodegenerative disorder, history of stroke, inflammatory disorders affecting the central nervous system (such as multiple sclerosis, encephalitis, meningitis, vasculitis), and inability to walk without aids. The study has been performed at the Department of Neurology and the Department of Psychiatry of the University Hospital Tuebingen, Germany. A large assessment battery with mainly quantitative, unobtrusive measurements for repeated objective application was designed. To avoid bias in data acquisition all investigators were blinded to the results of all other examinations. For more details about the TREND study please visit https://www.trend-studie.de/. Supplementary Figure 1 offers a flow chart of the exclusion criteria leading to the final dataset as an overview.

### Clinical investigations

#### Motor function: Gait

Gait assessments were performed in an at least 1.5 meters wide corridor allowing obstacle-free 20 meter walking. All subjects performed four single task conditions: 1. walking with habitual speed, 2. walking with maximum speed, 3. checking boxes with maximum speed while standing, and 4. subtracting serial 7s with maximum speed while standing. Additionally, two dual task conditions were performed: 1. walking with maximum speed and checking boxes with maximum speed. 2. walking with maximum speed and subtracting serial 7s with maximum speed ^7^.

Based on the two dual task conditions, we extracted the respective four dual-task speeds: 1. checking boxes when walking (number of boxes per second) 2. walking when checking boxes (meters per seconds) 3. subtracting when walking (number of serial 7s subtractions per second) 4. walking when subtracting (meters per second). The single and dual task speed parameters were then used to calculate dual task costs and overall speed according to the following formulae:

- Overall speed: dual-task-speed + single-task-speed
- Dual task cost: dual-task-speed – single-task-speed

#### Cognition

Detailed cognitive function was tested using the standardized German version of the extended Consortium to Establish a Registry for Alzheimer’s Disease (CERAD)-Plus neuropsychological battery ^8,9^. This comprehensive battery includes the following cognitive subtests: semantic and phonematic verbal fluency tasks, Boston Naming Test, Mini Mental Status Examination (MMSE), word list learning, word list recall, word list recognition, figure drawing, figure recall and the Trail Making Test (TMT) A and B^10,11^. The TMT consists of two parts and evaluates executive function, cognitive flexibility and working memory ^12^. In part A, participants connect randomly spread numbers from 1 to 25 in ascending order. In part B, participants are asked to connect randomly spread numbers (1 to 13) and letters (A to L) in alternating numeric and alphabetical order (1-A-2-B-3-C-…-13-L). In case of an error, the examiner draws the attention of the participant to the error, to allow completion of the task without errors at the expense of additional time. The maximum time allowed is 180 seconds for part A and 300 seconds for part B. After this time the investigator discontinues the experiment. Two parameters were calculated from the TMT A and TMT B test:

- Overall speed: TMT A + TMT B
- Cognitive flexibility: TMT B – TMT A

Next to the CERAD total score, subscores of the different CERAD domains were included in the analysis. Ordinal variables were measured on a Likert scale and indicate the number of items completed correctly.

### Medical Condition

#### Hypertension

Lifetime diagnosis of hypertension (medical history) and/or intake of anti-hypertensive medication was defined as presence of hypertension.

#### Obesity

Body mass index (BMI) was calculated by: mass [kg] / (height [m])^2^).

#### Body composition (fat/skeleton muscle mass)

was assessed by bioelectrical impedance analysis (BIA) using a body impedance analyser (BIA 101, Akern, Germany) for two out of four visits. Therefore, ohmic resistance was measured between the dominant hand wrist and dorsum and the dominant foot angle and dorsum in supine position. Muscle mass in kg was then calculated according to Janssen et al. ^13^ and subsequently normalized to subjects’ body height squared (skeletal muscle index: SMIBIA): with body height in centimeter, resistance in Ω, for gender: male = 1 and female = 0 and age in years.

### Lifestyle

#### Assessment of physical activity

Physical activity was assessed by a self-administered questionnaire. This questionnaire is part of the Bundes-Gesundheits Survey (national health survey) and allows to rate physical activity between 0 and 4 (0 = no activity, 1 = 0.5-1h per week, 2 = 1-2h per week, 3 = 2-4 hours per week, 4 = more than 4 hours per week) ^14^.

#### Smoking and drinking

Personal history of smoking and alcohol drinking behavior were assessed by a self-administered questionnaire. Pack-years were calculated by quantifying the packs (20 cigarettes/pack) smoked per day multiplied by years as a smoker. The frequency of drinking alcohol was assessed on a scale from 0-4 which indicates the number of drinks per month.

### Genetic risk factors for PD and AD

DNA was isolated from EDTA blood by salting out and stored at 4°C. All participants were analyzed by NeuroChip. Pathogenic variants in the most common PD-associated genes *LRRK2* and *GBA* were confirmed by Sanger sequencing. None of the participants carried a *LRRK2* mutation. Fifty-seven participants carried a *GBA* variant. We further grouped those according to known PD-specific mutation severity: wildtype (0), low risk (1) and mild/severe (2). Moreover, the most relevant Single Nucleotide Polymorphisms (SNPs) in genes for PD (*SNCA* rs356220 or proxy rs356219) and AD (*ApoE, MAPT*) were investigated to explore the effect on motor and cognitive function. We grouped the number of risk alleles according to an additive model: *SNCA* rs356220 (or proxy rs356219) minor allele C (0, 1, 2), *ApoE4* allele (0, 1, 2), and *MAPT* haplotype (H1/H1, H1/H2, H2/H2).

### Measurement of neurofilament light chain (NFL) in blood

Blood samples were collected at the day of the study visit, cooled, centrifuged (4°C, 10 minutes, 2000g), aliquoted, and stored at –80^?^C within 4 hours after collection. They were analyzed without any previous thaw–freeze cycle. Serum levels of NFL as marker for neuronal-axonal damage were measured in duplicates using the SIMOA NF-light KIT (Quanterix, Product number: 103186) on the SIMOA HD-1 Analyzer (Quanterix, Lexington, MA) as established previously^15^. Technicians were blinded to all other tests of the participants.

### Definition of age-related key functions

The aim was to simultaneously predict aging-related key functions of motor and cognitive performance from a large set of multiple multi-modal factors including genetic, biofluid, clinical, demographic and lifestyle factors (Supplementary Table 1).

As outcome measures for motor function we defined the different gait conditions:

- Overall speed: dual-task speed + single-task speed

∘ Walk while Subtract serial 7s Dual + Single Walk
∘ Subtract serial 7s while Walk Dual + Single Subtract serial 7s
∘ Walk while Cross boxes Dual + Single Walk
∘ Cross boxes while Walk Dual + Single Cross boxes
- Dual task cost: dual-task speed – single-task speed

∘ Walk while Subtract serial 7s Dual - Single Walk
∘ Subtract serial 7s while Walk Dual - Single Subtract serial 7s
∘ Walk while Cross boxes Dual - Single Walk
∘ Cross boxes while Walk Dual - Single Cross boxes

As outcome measures for cognitive function we used the CERAD and defined the different CERAD subdomains:

- Overall cognitive function: Total score
- Memory function: Word list learning and Word list recall
- Executive function: TMT A + B and TMT B - A

All scores were transformed such that higher values reflect worse performance by flipping the scale.

### Models Overview

We developed a Bayesian Reduced Rank Regression (RRR; see below for details) model that predicts all outcome measures from a single composite score extracted as a linear combination of all input factors (Supplementary Table 1). This restricts the flexibility of the model but increases its ability to identify a key feature extractor by using several prediction targets. We compared the predictive performance of the Bayesian RRR model to a standard approach with classical multivariate linear regression where different linear combinations can predict the outcome measures. Figure 1 illustrates the rationale for Bayesian Reduced Rank Regression compared to classical multivariate linear regressions.

We used python 3 (3.8.2) in combination with the probabilistic modeling library PyMC3 (3.9.3) ^16^ to implement the Bayesian RRR model. The OLS models were implemented with statsmodels (0.11.1) ^17^. Model evaluation was performed using scikit-learn (0.23.1) ^18^. We used datajoint (0.12.6) ^19^ to build our data processing pipeline.

### Handling Missing Data

Missing data of predictor variables were handled in the same way for both models. After subject and visit exclusion as detailed in Supplementary Figure 1 we assessed the missingness of the predictors across all remaining visits. The percentage of missingness for time-varying predictor variables can be found in Supplementary Table 1. To increase the amount of available data points we performed imputation. We did so only for subjects with at least one value available for each predictor. Based on the assumption that predictors only change when a new value is given, we first applied forward filling and then backward filling.

### Model Comparison

To compare the predictive performance of the Bayesian RRR model against a more flexible and traditional approach, we trained 13 ordinary least squares (OLS) models that each predict a single outcome measure from the set of predictors. To handle the longitudinal data, we decided to include all available visits of each subject; thereby, having subjects unequally represented in the dataset. The models thus treat each visit as an independent data point disregarding the correlation arising from repeated measures of the same subject. As the outcome measures have variable availability (Supplementary Table 2) for each model the valid visits to include for training were selected separately in order to maximize the number of overall data points. For each outcome measure we kept all data points where the outcome measure itself was available. We thus trained the OLS models on differently sized datasets. In contrast, the Bayesian RRR model was trained on all targets simultaneously. Due to the nature of the Bayesian framework, we can include data points where parts of the outcome measures are missing. Thereby the entire Bayesian RRR is trained on the union of the datasets for the OLS models but for each outcome measure only the same visits as for the corresponding OLS model are used for training. For the OLS models, we decided to use dummy encoding for nominal and ordinal predictors and include an intercept term. For a predictor with *n* categories, we thus included *n* − *1* coefficient into the model. Real-valued predictors were standardized. All models were fit using mean-squared error loss. We performed 5-fold cross validation using 20% as test and 80% as training data ensuring that visits of the same subject are grouped into either one. For each fold the outcome measures and real-valued predictors were standardized on the training set. As no hyperparameters (values that we set to control the learning process) were learned, cross validation yielded a measure of uncertainty for the prediction performance from the 5 folds. All performance evaluation and comparison was done through this cross-validation. We retrained all 13 OLS models on their respective complete dataset (training and test) to obtain the final coefficients for the predictors.

### Reduced Rank Regression Model (RRR)

Our reduced rank regression model is based on the observation that the outcome measures/clinical tests are correlated and thus can be represented through a smaller set of latent variables. Therefore, we employed an RRR model that allows us to predict multiple response variables from the same set of predictor variables while reducing the amount of model parameters (see Figure 1). RRR can be seen as a multivariate regression model with a coefficient matrix of reduced rank ^20^. RRR is a computationally efficient method which increases statistical power in settings where the number of dimensions is large compared to the number of examples. In such *m* ≫ *j* settings RRR is nowadays a state-of-the-art method in fields with high dimensional data such as genetics and imaging ^21,22^. Given *j* observations of *m* predictors and *n* outcome measures, standard multivariate regression requires fitting *m* · *n* coefficients *Y* = *XC* + *E*, with *Y* being the response matrix of size *j* × *n, X* being the *j* × *m* predictor matrix, *C* the *m* × *n* coefficient matrix and *E* being the error term matrix of size *j* × *n*. The RRR is obtained by adding a rank constraint *rank*(*C*) = *k, k* ≤ *min*(*n, m*). The rank constraint decreases the dimensionality of the model, and improves the statistical power. Using the rank constraint, *C* can be rewritten as *C* = *AB*^*T*^, with *A* of size *m* × *k* and *B* having size *n* × *k*. Hence, the model can be expressed as *Y* = (*XA*)*B*^*T*^ + *E*.

This decomposition allows for interpretations of *A* and *B. A* is a mapping from the predictor matrix *X* to a latent representation of dimension *k. B* is a mapping from the latent scores to the responses *Y*. The latent scores *XA* display the low-dimensional predictor variability that is predictive of the response variability.

### Bayesian Reduced Rank Regression

We used Bayesian inference for our RRR model to obtain parameter uncertainty and handle missing data. This means that given observations *X* and *Y*, we sampled model parameters *Ψ* from the posterior distribution *p*(*Ψ*|*X, Y*) ∝ *p*(*Y*|*Ψ, X*) · *p*(*Ψ*). The variable *Ψ* generically denotes all parameters of the model. The Bayesian framework requires a prior distribution *p*(*Ψ*) that embodies our prior knowledge about these parameters and the behavior that we want the model to exhibit. We are specifying these choices in the following paragraphs.

Least Squares Regression models can easily be transformed into Bayesian models by rewriting the model as *Y* ∼ *N*(*XAB*^*T*^, *σ*^*2*^), where *N*(*μ, Σ*) denotes the normal distribution with mean *μ* and covariance *Σ*

Given a high dimensional data setting it is likely that some of the predictors are non-informative for some of the outcome measures. A Laplace prior on *A* can realize this desired sparsity as it promotes element-wise sparsity: Certain elements in *A* are set to *0*, resulting in a latent composite score depending on certain predictors but not on others.

Suppose we have a predictor matrix *X* which holds information about *m* predictors (time-varying and static) for *j* visits of subjects. Through *A* those are mapped to the latent space of size *k*, such that we obtain *k* composite scores for each visit, *θ*. For visit *i* we thus get for composite score 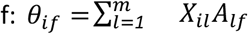. A priori, each element of *A* is sampled from a Laplace distribution 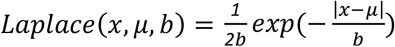 with *b* = *1* to enforce element-wise sparsity. The matrix *B* maps back from the latent space to the response space. A Lognormal distribution *ln*(*e* ^*μ* +,*σZ*^) with *σ* = *0*.*25* is used as a prior to enforce the positivity of the coefficients. By centering the real responses prior to learning, an offset can be omitted. For visit *i* we get a prediction for the response *o* via *Y*_*io*_ = *θ’*_*io*_ *B*_*o*_for which we assume a Gaussian observation noise with *σ* = *0*.*908* which is informed by the MSE on the training data of the multiple OLS models. We trained a model with *k* = *1* for which we present the results in the main text, but we also trained a model with *k* = *2* to check how this increase in complexity improves the performance. We further trained a model with *k* = *1* and a deterministic *B* = *1* to check whether our Bayesian model with similar complexity to the OLS models performs as well as those. The results can be found in Supplemental Table 3.

### Ordinal Predictors

We further improved our Bayesian RRR model through the way it handles ordinal predictors. Ordinal variables are commonly used in clinical settings. However, in most modeling approaches they are encoded as either nominal or interval variables. The former disregards the ordering information, and the latter assumes regular spacing which may not be given. To correctly use ordinal predictors, one can use monotonic effects ^23^ (see Supplementary Figure 2). This transformation ensures a monotonic increase or decrease while adjacent categories can be arbitrarily spaced. For an ordinal predictor *x* taking values *x*_*n*_ ∈ {*0*, …, *D*} a monotonic transformation is defined 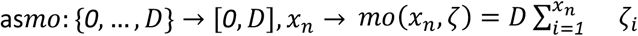 where 𝜁 is element of a simplex, meaning it satisfies 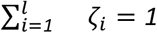 and 𝜁_*i*_ ∈ [*0,1*]. It can be interpreted as the normalized distances between adjacent categories. As D can be absorbed into the regression coefficients A and lead to redundancies, we instead encoded ordinal variables in our model with: 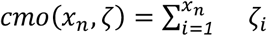.

This still ensures a monotonic transformation with arbitrary spacing, however the effect and sign will be inferred through the regression coefficient. In our Bayesian RRR model we chose a Dirichlet prior for the 𝜁_*i*_ as it is the natural choice for a prior on simplex parameters. By choosing a constant *α* = *1*, we effectively used a uniform (equal probability) prior on the probability simplex, i.e. all vectors 𝜁 that sum to one are equally likely. The a priori expectation of 𝜁 is given by 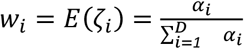. With *α* = *1* we have 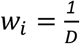. This prior centers the category distances, 𝜁, around a linear trend but allows for high variations around this. This transformation was applied to the ordinal predictors in *X* prior to the RRR.

We decided to model the genetic data as ordinal predictors as well. The monotonic transformation allows us to consider dominant (0 vs. 1), additive (0 vs. 1 vs. 2) as well as recessive (0 vs. 2) effects simultaneously.

### Sampling from the posterior

We sampled from the posterior *p*(*Ψ*|*X, Y*) through NUTS sampling ^24^ with two chains, each with a burn-in of 2000 samples and 500 retained samples. We thus obtained 1000 samples from the posterior distributions of each parameter. Subsequently, we obtained the posterior predictive distribution by feeding the samples through the generative model: *p*(*Y*|*X*) = ∫ *p*(*Y*|*Ψ*)*p*(*Ψ*|*X*)*dΨ*.

These predictions were used for performance evaluation. To assess the generalization of our model, we performed 5-fold cross validation where for each split the data was randomly split into a test (20%) and train (80%) set ensuring that each subject is only in either one. We retrained the model using the whole dataset to obtain the final posterior distributions for the coefficients.

### Performance Evaluation

We used the coefficient of determination R^2^ to compare our model performances. Suppose we have *n* data points with *y*_*i*_ being the true value for visit *i, ŷ*_*i*_ being our predicted value, and *ŷ*− being the mean of the true values. R^2^ is defined as 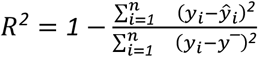.

It is *1* for perfect prediction and *0* when the mean is predicted. Note, that R2 can be negative if the prediction is worse than the mean, i.e. the constant predictor. We calculated R^2^ as the standardized mean-squared error (MSE): 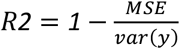 where y are the true values. To make the measure more robust, we decided to normalize the MSE by the variance of the whole dataset, i.e. train and test set. This better captures the true variation regardless of the applied train/test split. We compared the performance over the 5 folds of the OLS models and the Bayesian RRR model for each clinical test with a t-test.

We evaluated the significance of predictors of the multiple linear regression models with t-tests and a type-I error threshold for the p-value of 0.05 that is corrected by the number of tests performed using Bonferroni correction. For our Bayesian RRR model where we obtained posterior samples for our coefficients, we calculated the highest posterior density ^25^ (95%) and defined significance as this interval not crossing 0.

### Ethics

The study was approved by the Ethics Committee of the Faculty of Medicine at the University of Tübingen (TREND: 90/2009BO2), and all participants gave written informed consent.

## Data Availability

Anonymized data can be shared upon request. Study data were collected and managed using REDCap electronic data capture tools hosted at University of Tuebingen.

## Data availability

Anonymized data can be shared upon request. Study data were collected and managed using REDCap electronic data capture tools hosted at University of Tübingen^26^.

## Author Contributions

DB, KB, WM, and GE designed the TREND study. All authors were involved in data collection or processing. AS and FS designed the model and performed the statistical analysis. AS implemented the code for all models and analyses. AS, SL, KB, FS drafted the manuscript. All authors were involved in interpretation of the data and critical revision of the manuscript. All authors gave their final approval.

