## Supplements for "Machine learning-based personalized composite score dissects risk and protective factors for cognitive and motor function in elderly"

### Supplementary material

**Supplementary Figure 1: Data selection flowchart. The decision flowchart is depicted which details the exclusion criteria and amount of subjects/visits lost due to those selections.**

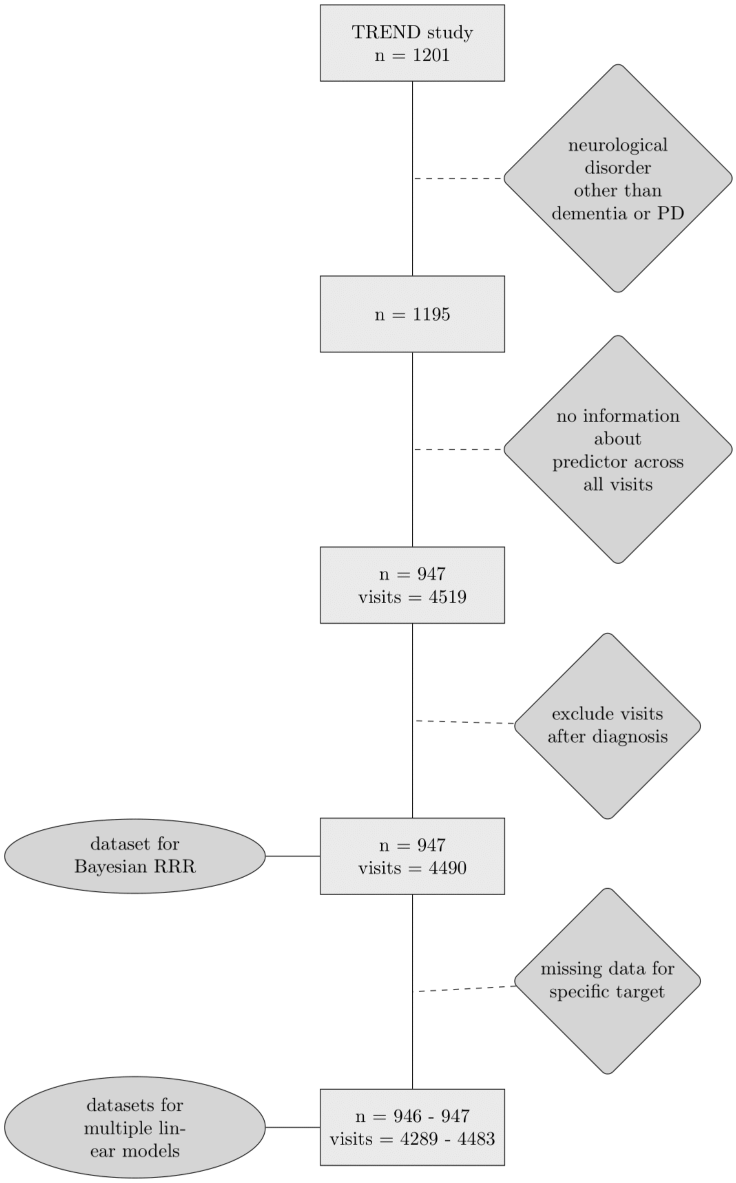

**Supplementary Figure 2: Schematic of monotonic transformation.** *An ordinal variable with 4 categories (x-axis) is shown with the learned monotonic transformation (y-axis).* $\zeta_i$ *depicts the learned distance between two categories and their sum (distance between first and last category) equals 1.*

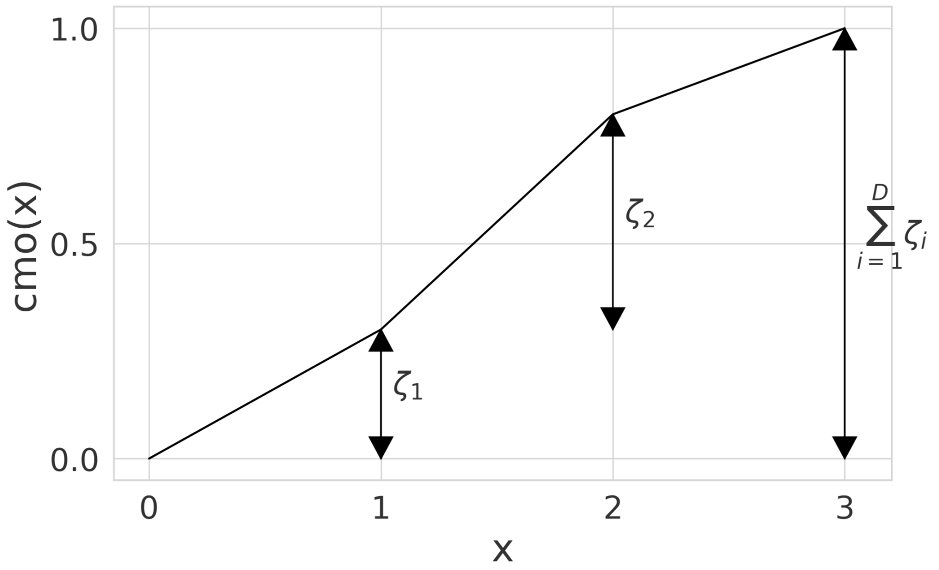

**Supplementary Figure 3: The two composite scores model recovers the opposite effect of factors on cognitive and gait-related outcome measures.** *The color indicates the direction and size of the effect of a predictor (x-axis) onto a target (y-axis). The size of the square indicates its importance as the absolute ratio of mean and standard deviation (the larger the further away from 0).*

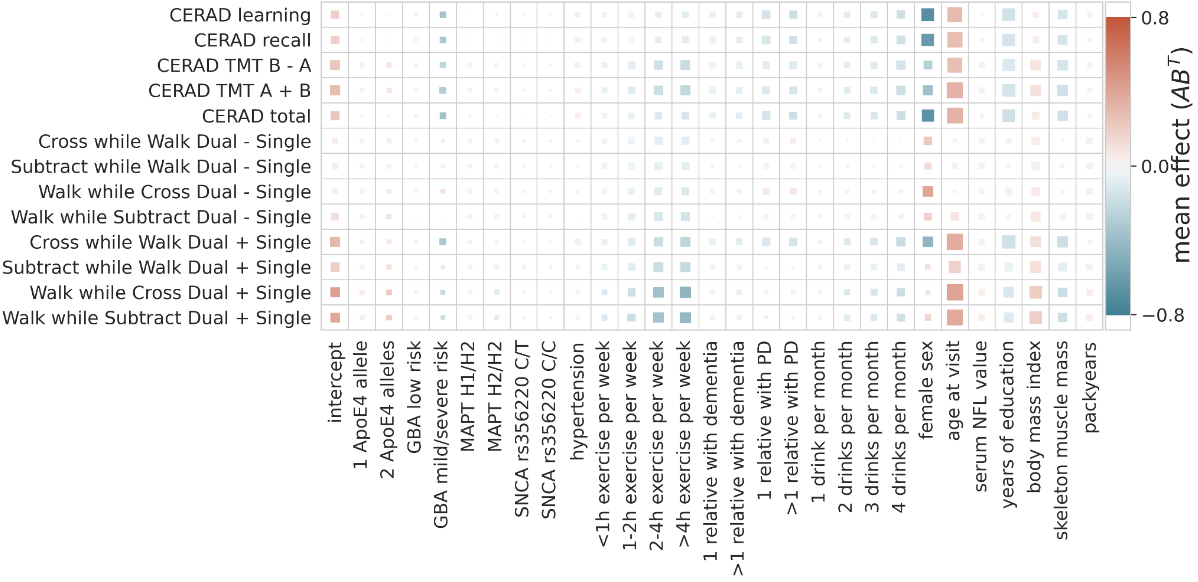
CERAD learning
CERAD recall
CERAD TMT B - A
CERAD TMT A + B
CERAD total
Cross while Walk Dual - Single
Subtract while Walk Dual - Single
Walk while Cross Dual - Single
Walk while Subtract Dual - Single
Cross while Walk Dual + Single
Subtract while Walk Dual + Single
Walk while Cross Dual + Single
Walk while Subtract Dual + Single
intercept
1 ApoE4 allele
2 ApoE4 alleles
GBA low risk
GBA mild/severe risk
MAPT H1/H2
MAPT H2/H2
SNCA rs356220 C/T
SNCA rs356220 C/C
hypertension
<1h exercise per week
1-2h exercise per week
2-4h exercise per week
>4h exercise per week
1 relative with dementia
>1 relative with dementia
1 relative with PD
>1 relative with PD
1 drink per month
2 drinks per month
3 drinks per month
4 drinks per month
female sex
age at visit
serum NFL value
years of education
body mass index
skeleton muscle mass
packyears
0.8
0.0
−0.8
mean effect ($AB^T$)

**Supplementary Figure 4: Regression coefficients for the two composite scores model.** *The estimated effect sizes of the predictors on the composite score are displayed. The highest posterior density is plotted. The coloring indicates significance (95% highest posterior density contains 0, not significant = gray) and direction of the effect (blue = protective, red = risk).*

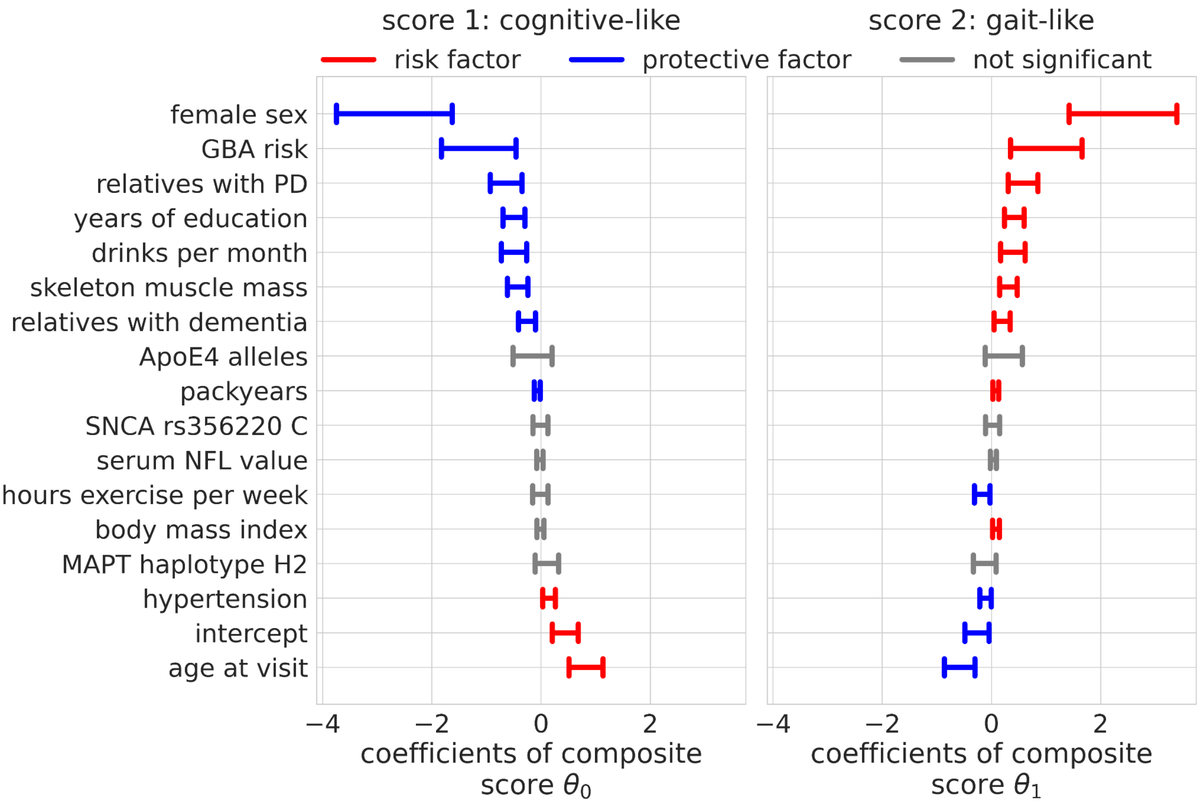

**Supplementary** ***Table 1: Statistics of predictor variables/factors.*** *The mean/median of the selected predictor variables (and conversion to diagnosis) are shown along with their standard deviation/quartiles. The % missing from the union of all visits (4490 datasets for 947 subjects) is shown. These values were imputed. *This data was only collected at two out of four visits per study protocol.*

| normal/categorical data | % missing | mean | std |
|---|---|---|---|
| age at visit | 0 | 66.828 | 7.131 |
| serum NFL | 19.065 | 16.595 | 16.595 |
| years of education | 0 | 14.443 | 2.665 |
| female sex | 0 | 0.487 | 0.500 |
| converts to PD during study | 0 | 0.015 | 0.122 |
| converts to dementia during study | 0 | 0.018 | 0.135 |
| body mass index | 0.646 | 26.112 | 4.224 |
| skeleton muscle mass | 52.183 * | 26.091 | 6.678 |
| packyears | 7.572 | 6.214 | 12.232 |
| hypertension | 0 | 0.611 | 0.488 |
| ordinal data | % missing | median | [25%, 75%] |
| ApoE4 alleles | 0 | 0 | [0,0] |
| MAPT haplotype H2 | 0 | 0 | [0,1] |
| GBA pd risk | 0 | 0 | [0,0] |

| | | | |
|---|---|---|---|
| SNCA rs356220 C | 0 | 1 | [*1*,*2*] |
| hours exercise per week | *21.114* | 3 | [*2*,*3*] |
| drinks per month | *33.185* | 4 | [*2*,*4*] |
| relatives with PD | 0 | 0 | [*0*,*0*] |
| relatives with dementia | 0 | 0 | [*0*,*1*] |

**Supplementary Table 2: Statistics of outcome measures.** The number of data points (count), mean/median and quartiles/standard deviation are shown for each of the included clinical tests.

| ordinal data | count | median | [*25%*, *75%*] |
|---|---|---|---|
| CERAD total | 4472 | 14 | [*9*,*20*] |
| CERAD learning | 4399 | 9 | [*7*,*12*] |
| CERAD recall | 4483 | 3 | [*2*,*5*] |
| **normal data** | **count** | **mean** | **std** |
| CERAD TMT B - A | 4425 | *51.170* | *34.758* |
| CERAD TMT A + B | 4425 | *126.188* | *48.771* |
| Cross while Walk Dual - Single | 4383 | *−0.279* | *0.264* |
| Subtract while Walk Dual - Single | 4289 | *−0.010* | *0.114* |
| Walk while Cross Dual - Single | 4399 | *−0.219* | *0.166* |
| Walk while Subtract Dual - Single | 4312 | *−0.298* | *0.207* |
| Cross while Walk Dual + Single | 4383 | *−2.727* | *0.521* |
| Subtract while Walk Dual + Single | 4289 | *−0.679* | *0.292* |

| | | | |
|---|---|---|---|
| Walk while Cross Dual + Single | 4399 | *−3.037* | *0.463* |
| Walk while Subtract Dual + Single | 4312 | *−2.962* | *0.453* |

**Supplementary Table 3: Results of t-test for significance of performance difference.** We compare the mean performance of each of the Bayesian models (composite score: k=1, two composite scores: k=2) over 5 folds to the OLS models' performance over 5 folds with a t-test. The test statistic t along with the p-value is shown, where a p-value smaller than 0.05 indicates significance.

| k | 1 | | 2 | | single task | |
|---|---|---|---|---|---|---|
| test | t-statistic | p-value | t-statistic | p-value | t-statistic | p-value |
| CERAD learning | -0.771582 | 0.462542 | 0.903303 | 0.392751 | 1.18038 | 0.271753 |
| CERAD recall | 0.390882 | 0.706088 | 1.447446 | 0.185797 | 1.59984 | 0.148302 |
| CERAD TMT B - A | 0.761247 | 0.468353 | 0.274502 | 0.790655 | 0.726159 | 0.488439 |
| CERAD TMT A + B | 0.579235 | 0.578372 | -0.382270 | 0.712216 | 1.14434 | 0.285564 |
| CERAD total | -0.624379 | 0.549766 | -0.097527 | 0.924707 | 1.2544 | 0.245106 |
| Cross while Walk Dual - Single | -2.136732 | 0.065107 | -0.979398 | 0.356069 | -1.16826 | 0.276338 |
| Subtract | -2.978478 | 0.017641 | -1.003217 | 0.345132 | -0.740939 | 0.479911 |

| | | | | | | |
|---|---|---|---|---|---|---|
| while Walk Dual - Single | | | | | | |
| Walk while Cross Dual - Single | -3.184523 | 0.012909 | -0.341609 | 0.741445 | -0.353704 | 0.732701 |
| Walk while Subtract Dual - Single | -0.722037 | 0.490835 | 0.060063 | 0.953579 | -0.193342 | 0.85151 |
| Cross while Walk Dual + Single | -2.050907 | 0.074403 | -2.874170 | 0.020696 | -1.22074 | 0.25694 |
| Subtract while Walk Dual + Single | 0.445314 | 0.667896 | 0.179760 | 0.861811 | 0.584819 | 0.574788 |
| Walk while Cross Dual + Single | -2.717913 | 0.026334 | -4.846756 | 0.001277 | -0.332764 | 0.747865 |
| Walk while Subtract Dual + | -2.956444 | 0.018245 | -5.664771 | 0.000473 | -0.198769 | 0.847402 |

| |
|---|
| Single |
